# Short-term psychosocial outcomes following disclosure of glaucoma polygenic risk score, INSiGHT Study

**DOI:** 10.64898/2026.06.24.26356219

**Authors:** Giorgina Maxwell, Robert Allen, Lucinda Hodge, Simone Kelley, Jamie E Craig, Sarah Cohen-Woods, Emmanuelle Souzeau

**Affiliations:** Department of Ophthalmology, Flinders Medical and Health Research Institute, Flinders University, Adelaide, SA, Australia; Department of Psychology, Flinders University Institute for Mental Health and Wellbeing, Flinders University, SA, Australia

**Keywords:** Glaucoma, polygenic risk score, genetic risk stratification, genetic testing

## Abstract

Early glaucoma detection and treatment are critical to prevent irreversible blindness. Glaucoma polygenic risk scores (PRS) offer an effective approach for stratifying disease risk and are increasingly available in clinical practice. However, the psychosocial impact of receiving glaucoma PRS results is currently unknown. As such, this study investigated short-term psychosocial outcomes of disclosing glaucoma PRS to individuals over 50 years from the general population. Individuals from the bottom 10%, middle 45-55%, and top 10% of PRS scores were invited to receive their results and complete surveys before and 2 weeks after receiving results to assess anxiety, test-related distress, decisional regret, recall and understanding.

Of invited participants, 51.7% (136/263) enrolled with 133 completing both surveys. Two weeks after disclosure, PRS recall was high (78.2%), although PRS knowledge remained limited. Privacy concerns were moderate, not differing across PRS groups (*Χ^2^* = 4.17, *p* = .124). Small reductions in glaucoma-related anxiety (*z* = –2.93, *p* = .003), generalised anxiety (*z* = −3.75, *p* < .001) and stress (*z* = −2.49, *p* = .013) were observed following disclosure. While scores remained within normal ranges, higher glaucoma-related anxiety (*t* = –2.36, *p* = .020), higher negative emotions (*Χ^2^* = 20.80, p < .001), and lower positive experience (*F* = 5.70, *p* = .004) were seen for high-risk participants compared to lower-risk participants. Decisional regret was low and didn’t differ across PRS groups (*Χ^2^* = 0.28, *p* = .869). These findings support the psychosocial safety of glaucoma PRS testing while highlighting the need for improved education and longer-term follow-up to support clinical implementation.

## Introduction

Despite diagnostic and therapeutic advancements, glaucoma remains the leading cause of irreversible blindness globally, and is predicted to affect 111.8 million by 2040 (1,2). Evidence-based treatment can minimise progression but cannot restore lost vision; therefore, early diagnosis and intervention are critical (3). However, glaucoma is frequently undiagnosed, with an estimated half of affected individuals unaware they have the disease (4). Factors contributing to current underdiagnosis include the asymptomatic nature of early disease and the lack of cost-effective, widely supported population screening programs, highlighting the need for better strategies to identify at-risk individuals early to prevent vision loss (5).

The genetic heritability of adult-onset primary open-angle glaucoma (the most common type of glaucoma; subsequently referred to as ‘glaucoma’) is estimated at 70%, making it one of the most highly heritable complex diseases (6). Current polygenic risk scores (PRS) for glaucoma account for 9.4-14.9% of its heritability (7,8), and are showing improved performance across ancestry groups (7,9). Glaucoma PRSs have some of the highest effect sizes among complex diseases, including cardiovascular disease and inherited cancers (10). Individuals in the top 10% of the PRS distribution have a 10-fold increased risk relative to those in the remaining 90% (11). Clinically, a higher glaucoma PRS correlates with greater disease risk, earlier onset, stronger family history, more aggressive disease progression, a greater likelihood of requiring surgical intervention, and faster vision loss despite treatment (12–16), supporting its role in guiding screening and interventions (17).

PRS testing for glaucoma is currently clinically accredited and available in countries including Australia, New Zealand and the United States. Despite growing commercial availability, gaps in the literature remain regarding effective clinical implementation and uptake. Our previous research supports interest and acceptability in the test from both patients and healthcare professionals (18,19). Research to date has shown limited long-lasting or impactful psychosocial harms following polygenic testing for inherited cancers or cardiac disease, although a minority of individuals report increased distress or disease-related worry (20). To date, no studies have investigated the psychosocial impact of polygenic testing in the context of glaucoma. As such, the aim of this study was to evaluate the short-term psychosocial outcomes of receiving PRS results for glaucoma in adults from the general population.

## Methods

### Study design and participants

This study has been approved by the Southern Adelaide Clinical Human Research Ethics Committee (SAC HREC, 2023/HRE0085); the study protocol has been described elsewhere (21). Participants were recruited from an established cohort, the GRADE (Genetic Risk Assessment of Degenerative Eye Disease) study, which aims to apply glaucoma PRS testing to 1,000 individuals ≥50 years from the general population, and clinically examine individuals across the PRS spectrum (22). Participants in GRADE were recruited via various approaches including individuals who previously completed questionnaires on the acceptability of glaucoma polygenic testing, advertisement through clinics, community groups, and volunteer services. Individuals already diagnosed with glaucoma were neither targeted nor excluded to reflect the general population. Additional eligibility criteria to the GRADE study included sufficient proficiency in English to be able to complete questionnaires.

### Recruitment and data collection

Individuals from the GRADE study were invited to receive their research glaucoma PRS results, calculated from Craig et al (12). 330 participants were invited to participate in this study including 116 from the lowest decile (10%), 102 from the middle range (45–55%), and 112 from the highest decile (10%) of the PRS distribution.

Eligible individuals were sent an invitation letter and study information by email or post. Participants indicated their interest by returning a form by post or responding by email. A member of the research team (R.A., L.H., S.K.) contacted them up to three times (email or phone). At enrolment, the potential implications of receiving genetic results, including possible psychosocial and life insurance implications were discussed. All participants provided written consent.

Participants who chose to receive their results completed a baseline questionnaire prior to receiving their results, and a follow-up questionnaires two weeks after receiving results. Questionnaires were distributed via email or post according to participant preference. One to two weeks after results were sent, a study team member called participants to answer any questions. Up to three reminders (phone or email) were provided for incomplete questionnaires. Participants received a personalised, patient-centred PRS report (low-, average-, or high-risk) by email or mail (23). To ensure equity in access, participants were not excluded based on ethnicity, however the PRS report stated it was best validated for European ancestries. As no clinical guidelines currently exist for glaucoma monitoring based on PRS results, reports included existing Australian recommendations for regular eye health checks from age 50 years

(24). As no accredited polygenic testing for glaucoma was available at study commencement, participants initially received their research PRS results. Following the introduction of accredited testing during the study, all results were confirmed using National Association of Testing Authorities (NATA)-accredited testing based on the same PRS construct and returned to participants at study completion. Data were collected using self-reported questionnaires in REDCap. Questionnaires returned by mail were checked for completeness and manually entered into REDCap. Participants were recontacted if responses were incomplete (R.A., L.H., S.K.).

### Measures

Sociodemographic characteristics and medical history included age, gender, ethnicity, level of education, employment status, language spoken at home, whether they had biological children, family history of glaucoma, and self-reported glaucoma status. Details of the surveys conducted are provided in the study protocol (21). Baseline and week 2 questionnaires included the Depression Anxiety and Stress Scale (DASS) (25) to measure generalised depression, anxiety, and stress; the Impact of Events Scale (IES) (26), adapted for glaucoma to measure glaucoma-specific anxiety; 12 true/false items assessing knowledge of glaucoma; and four items to assess Genetic Determinism. The Feelings About Genomic Testing Results (FACToR) questionnaire to measure test-related distress (27); the Decisional Regret Scale to measure decision satisfaction and conflict (28); Recall and interpretation of results; and Result expectation questions were assessed at week 2 only.

### Statistical analysis

Data were collected between July 2023 and January 2025 and analysed using SPSS (Version 31; IBM Corp., 2024). Descriptive statistics summarised sociodemographic, clinical, and psychological characteristics. Internal consistency of multi-item scales was assessed using McDonald’s Omega (Supplementary Table 1). Changes in IES and DASS-21 scores from baseline to week 2 were evaluated using Wilcoxon Signed-Ranks tests as the paired differences were skewed and could not be adequately normalised using standard transformations, violating the assumptions of the paired t-test. Quade nonparametric ANCOVAs were used to examine the influence of PRS group on week 2 IES and DASS-21 scores while controlling for baseline values as residuals were not normally distributed and the assumption of homoscedasticity was violated, precluding the use of Analysis of Covariance (ANCOVA). FACToR positive experience subscale scores were analysed using one-way ANOVA with Tukey post hoc tests. FACToR subscales, negative emotions, uncertainty, and privacy concerns, were assessed using the Kruskal-Wallis tests with Dunn’s pairwise comparisons and Bonferroni correction for multiple testing as data were not normally distributed and the assumption of equal variances was violated, precluding the use of one-way ANOVA.

The relationship between demographic factors psychosocial outcome scores (controlling for baseline measures where relevant) was assessed using Mann–Whitney U tests for sex and family history, and one-way ANOVA for education and age (Supplementary Table 5). The association between PRS group and week 2 psychosocial outcome scores, adjusting for baseline measures and demographic factors, was examined using Quade non-parametric ANCOVA due to violations of normality and homoscedasticity assumptions (Supplementary Table 6).

## Results

### Response rate

Of the 330 individuals invited to the study, 5 were deceased, 112 declined participation, 62 could not be reached, and 151 consented to participate. Of those who consented, 139 completed the baseline questionnaire, with 136 also completing the week 2 questionnaire. This yielded a participation rate of 51.7% (136/263), excluding those deceased or who could not be reached (Figure 1). Three participants stated they had not received their results by the time they completed the week 2 questionnaire and were excluded from analyses, providing a final number of 133 participants for whom data is analysed and reported in this paper.

**Figure 1.**
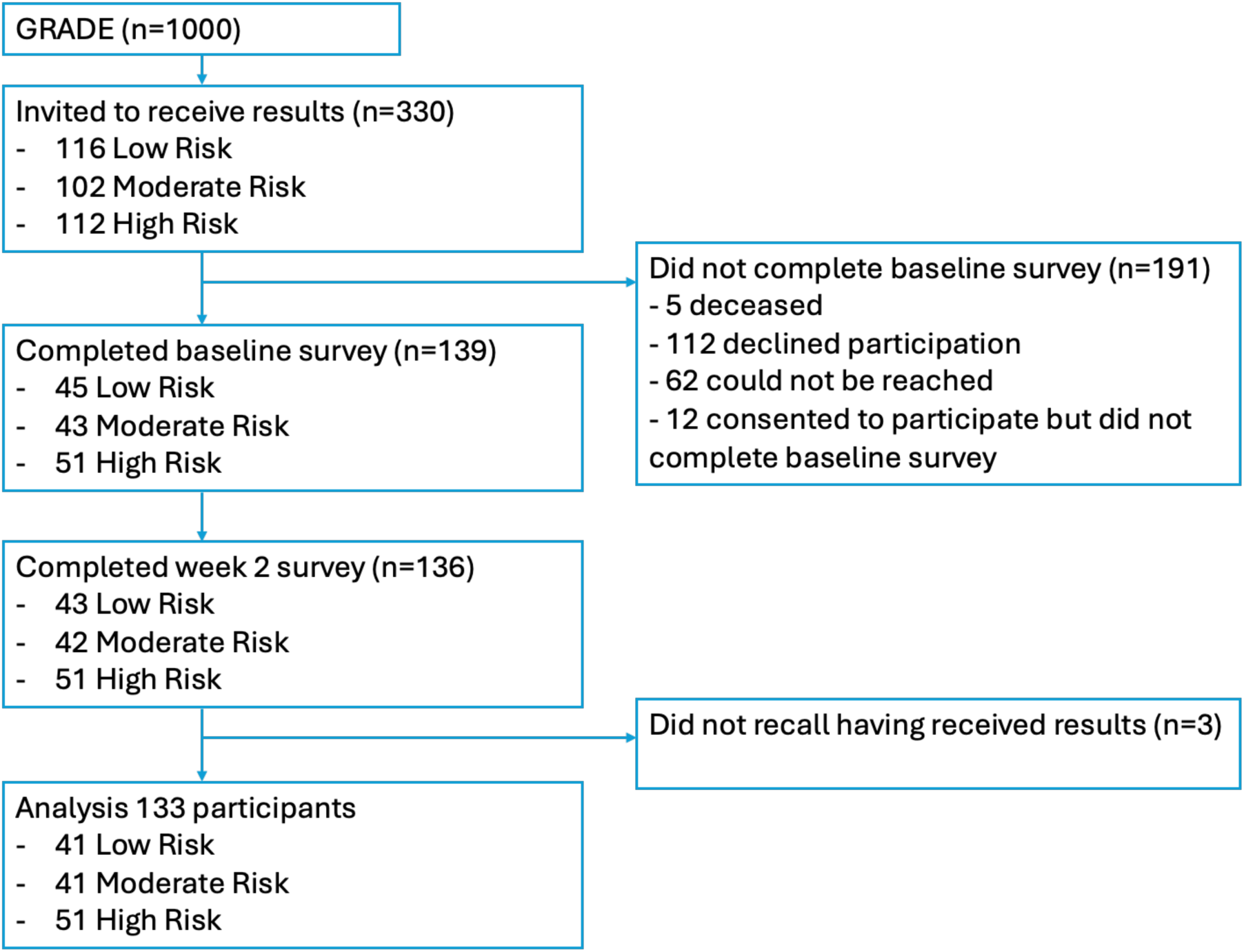
Participant recruitment flowchart.

Table 1 presents the characteristics of participants who completed the first survey, referred to as ‘Enrolled’, compared with those who did not, the ‘Declined’ (those who did not complete baseline survey less deceased). Those enrolled were on average younger than decliners and were more likely to be personally affected or have a family history of glaucoma.

**Table 1.** Enrolled and declined participant characteristics.

| Variable | Enrolled (n=139) | Declined (n=186) | Test statistic, p-value |
| --- | --- | --- | --- |
| Gender (Female) | 88 (63.3%) | 114 (61.0%) | $\chi^2 = 0.14$, .71 |
| Age (Mean (SD)) | 69.7 (8.7) | 71.9 (9.4) | $t = 2.16$, .03* |
| Ethnicity (Caucasian) | 126 (90.6%) | 164 (88.2%) | $\chi^2 = 0.51$, .48 |
| Glaucoma status (Affected) | 10 (7.2%) | 3 (1.6%) | $\chi^2 = 6.45$, .01* |
| Family history of glaucoma (Positive) | 48 (34.5%) | 34 (18.3%) | $\chi^2 = 11.14$, <.001*** |
| PRS<br>Low | 46 (33.1%) | 69 (36.9%) | $\chi^2 = 0.80$, .67 |
| Moderate | 43 (30.9%) | 59 (31.6%) |  |
| High | 50 (36.0%) | 59 (31.6%) |  |

Asterisks denote statistical significance: \* $p < 0.05$ ; \*\* $p < 0.01$ ; \*\*\* $p < 0.001$

Table 2 summarises the characteristics of the 133 participants included in the analysis. The majority identified as Australian or European (90.6%) and spoke English at home (98.6%). Self-reported glaucoma status and family history were more prevalent in individuals from the high-risk group, though not significantly associated (Table 2).

**Table 2.** Participant characteristics of those who completed Baseline and Week 2 survey.

| Variable | Low risk<br>(n=41) | Moderate risk<br>(n=41) | High risk<br>(n=51) | Test statistic, p-value |
| --- | --- | --- | --- | --- |
| Gender (Female) | 23 (56.1%) | 22 (53.7%) | 41 (80.4%) | $X^2 = 9.01, .01^*$ |
| Age (Mean (SD)) | 69.5 (9.53) | 69.1 (8.34) | 70.7 (8.30) | $F = .428, .65$ |
| Ethnicity (Caucasian) | 37 (90.2%) | 40 (97.6%) | 48 (94.1%) | $X^2 = 10.31, .24$ |
| Education | | | | $X^2 = 2.99, .56$ |
| University | 15 (36.6%) | 13 (31.7%) | 20 (39.2%) |  |
| Vocational | 17 (41.5%) | 14 (34.1%) | 14 (27.5%) |  |
| High school | 9 (22.0%) | 14 (34.1%) | 17 (33.3%) |  |
| Employment | | | | $X^2 = 1.78, .78$ |
| Employed | 8 (19.5%) | 12 (29.3%) | 12 (23.5%) |  |
| Retired | 30 (73.2%) | 25 (61.0%) | 36 (70.6%) |  |
| Other | 3 (7.3%) | 4 (9.8%) | 3 (5.9%) |  |
| Glaucoma status<br>(Affected) | 2 (4.9%) | 1 (2.4%) | 7 (13.7%) | $X^2 = 4.56, .09$ |
| Family history (Yes) | 12 (29.3%) | 11 (26.8%) | 24 (47.1%) | $X^2 = 5.03, .08$ |
| Biological children<br>(Yes) | 35 (85.4%) | 37 (90.2%) | 46 (90.2%) | $X^2 = 0.67, .72$ |

Asterisks denote statistical significance: \* $p < 0.05$ ; \*\* $p < 0.01$ ; \*\*\* $p < 0.001$

### Expectation of results

Overall, 48.9% of participants reported that their result fit with their family history of glaucoma, 13.5% reported it did not, and 37.6% were unsure (Supplementary Table 2). Participants’ expectations of whether their PRS aligned with their family history did not differ by PRS group (*X^2^* = 4.65, *p* = .325). In a multinomial logistic regression controlling for family history with “unsure” as the reference category, PRS groups were not associated with result expectation aligning or not aligning with family history (Supplementary Table 3).

### Recall of results

Two weeks after results disclosure, most participants (78.2%) were able to correctly recall their PRS category (higher, lower, or the same risk as the general population), 7.5% had forgotten or were unsure of their results, and 14.3% incorrectly recalled their results (Supplementary Table 2). There was no difference in ability to correctly recall PRS category across PRS groups (*X^2^* = 4.65, *p* = .325).

### Knowledge of glaucoma and PRS

Questions for the knowledge measure assessed different aspects of glaucoma and genetic risk understanding and as such internal consistency is not reported for this measure. Questions relating to genetic risk (Q1-8) were more commonly answered incorrectly while questions relating to glaucoma screening and management (Q9-12) were more commonly answered correctly (Supplementary Table 4). There was a small increase in knowledge post-result disclosure compared to baseline (*t* = 5.351, *p* < .001) but there was no significant difference in knowledge at week 2 according to PRS groups (*F* = 0.002, *p* = .998).

### Decisional regret

Decisional regret did not differ across assessed psychosocial and demographic factors (Supplementary Table 5). PRS was not significantly associated with decisional regret (*X^2^* = 0.28, *p* = .869), and this relationship was not changed when controlling for covariates (Supplementary Table 6).

### Depression, Anxiety, and Stress (DASS-21)

Internal consistency was good to excellent for depression and stress, and acceptable for baseline anxiety; however, week 2 anxiety showed lower reliability, suggesting that anxiety findings at this timepoint should be interpreted with caution (Supplementary Table 1). Average scores across the three subscales of the DASS were within the normal-to-mild range at baseline and week 2 (Table 3). Small reductions in subscale scores (anxiety, stress and depression) were observed in the full cohort post results disclosure (Table 3). DASS subscale scores were not associated with any of the assessed demographic or psychosocial factors (Supplementary Table 5).

**Table 3.** General anxiety and glaucoma-related anxiety at baseline and week 2.

|  | Baseline scores | Week 2 scores | Change over time<sup>1</sup> | PRS group effect at Week 2<sup>2</sup> |
| --- | --- | --- | --- | --- |
| Measure | Mean (SD) | Mean (SD) | z, p-value | F, p-value |
| **DASS – Depression (range: 0-42, normal scores: 0-4, mild scores: 5-6)** |  |  |  |  |
| All receivers | 4.3 (6.1) | 2.9 (4.4) | -3.20, .001** | 1.73, .182 |
| PRS (low) | 3.9 (4.7) | 3.3 (4.4) |  |  |
| PRS (moderate) | 3.8 (6.8) | 1.5 (2.3) |  |  |
| PRS (high) | 5.0 (6.7) | 3.8 (5.4) |  |  |
| **DASS – Anxiety (range: 0-42, normal scores: 0-3, mild scores: 4-5)** |  |  |  |  |
| All receivers | 3.1 (4.9) | 2.0 (2.9) | -3.75, <.001*** | 0.02, .983 |
| PRS (low) | 3.0 (3.9) | 1.8 (2.4) |  |  |
| PRS (moderate) | 2.5 (6.4) | 1.3 (2.1) |  |  |
| PRS (high) | 3.7 (4.3) | 2.6 (3.7) |  |  |
| **DASS – Stress (range: 0-42, normal scores: 0-7, mild scores: 8-9)** |  |  |  |  |
| All receivers | 5.9 (6.3) | 5.0 (5.7) | -2.49, .013* | 0.85, .431 |
| PRS (low) | 6.3 (6.8) | 6.1 (6.7) |  |  |
| PRS (moderate) | 5.7 (7.3) | 3.9 (4.1) |  |  |
| PRS (high) | 5.6 (5.1) | 4.9 (6.0) |  |  |
| **IES total (range: 0-75)** |  |  |  |  |
| All receivers | 6.5 (11.9) | 3.6 (8.2) | -2.93, .003** | 3.12, .048* |
| PRS (low) | 6.2 (10.3) | 3.5 (9.1) |  |  |
| PRS (moderate) | 5.8 (13.9) | 1.8 (5.1) |  |  |
| PRS (high) | 7.3 (11.5) | 5.1 (9.3) |  |  |
| **IES intrusion (range: 0-35)** |  |  |  |  |
| All receivers | 2.9 (5.6) | 1.4 (4.2) | -3.50, <.001*** | 0.55, .581 |
| PRS (low) | 2.9 (5.1) | 1.4 (4.5) |  |  |
| PRS (moderate) | 2.6 (6.5) | 0.8 (2.6) |  |  |
| PRS (high) | 3.1 (5.4) | 2.0 (4.9) |  |  |
| **IES avoidance (range: 0-40)** |  |  |  |  |
| All receivers | 3.6 (6.6) | 2.2 (4.3) | -2.25, .025* | 2.63, .076 |
| PRS (low) | 3.3 (5.6) | 2.1 (4.8) |  |  |
| PRS (moderate) | 3.2 (7.6) | 1.0 (2.6) |  |  |
| PRS (high) | 4.2 (6.5) | 3.1 (4.7) |  |  |

<sup>1</sup> Wilcoxon signed-rank test comparing baseline and week 2 scores across the cohort.
<sup>2</sup> ANCOVA examining differences in week 2 scores across PRS groups, controlling for baseline scores.
Asterisks denote statistical significance: \* p<0.05; \*\* p < 0.01; \*\*\* p<0.001

Controlling for baseline scores, week 2 DASS subscale scores were not associated with PRS group (Figure 2; Table 3), and this relationship was not changed when controlling for covariates (Supplementary Table 6).

**Figure 2.**
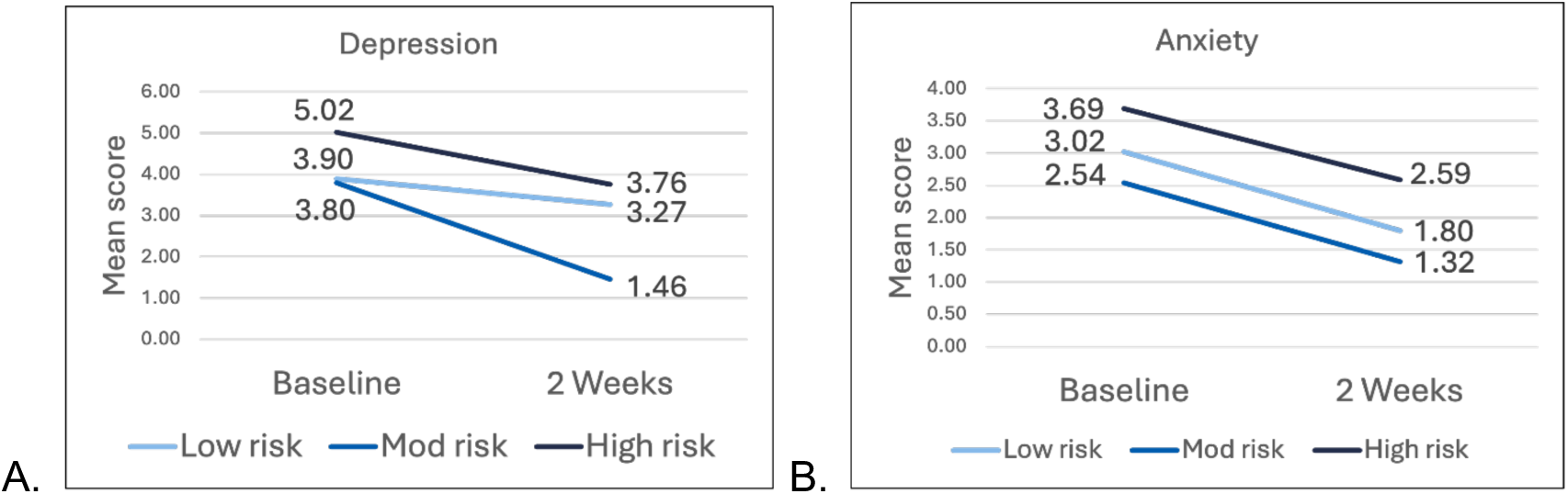
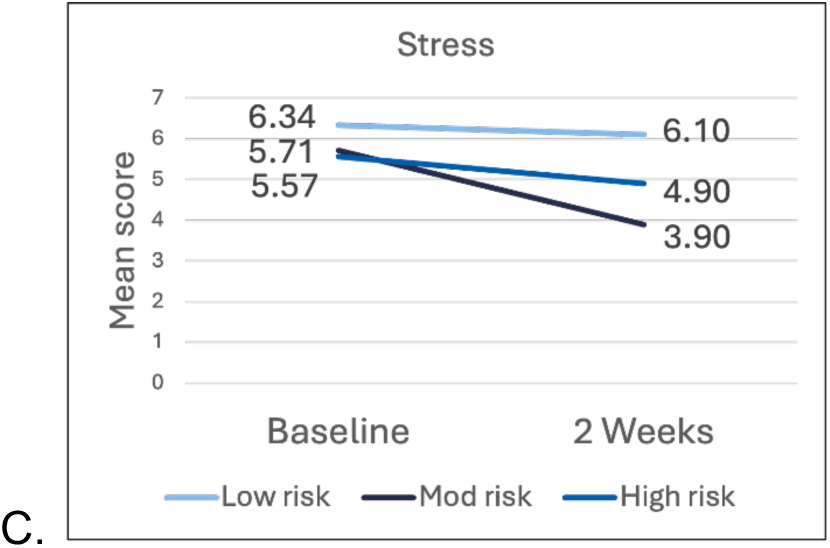
Comparison of mean DASS-21 scores measuring A) depression, B) anxiety and C) stress across PRS groups from Baseline to Week 2.

### Glaucoma-related anxiety (IES)

Glaucoma-related anxiety was assessed using total scores from the Impact of Event Scale, with additional analyses examining the intrusion and avoidance subscales. Average total and subscale IES scores were low at baseline and week 2 (Figure 3, Table 3). Small reductions in total and subscale scores were observed in the full cohort post results disclosure. IES total and subscale scores were not associated with any of the assessed demographic or psychosocial factors (Supplementary Table 5).

**Figure 3.**
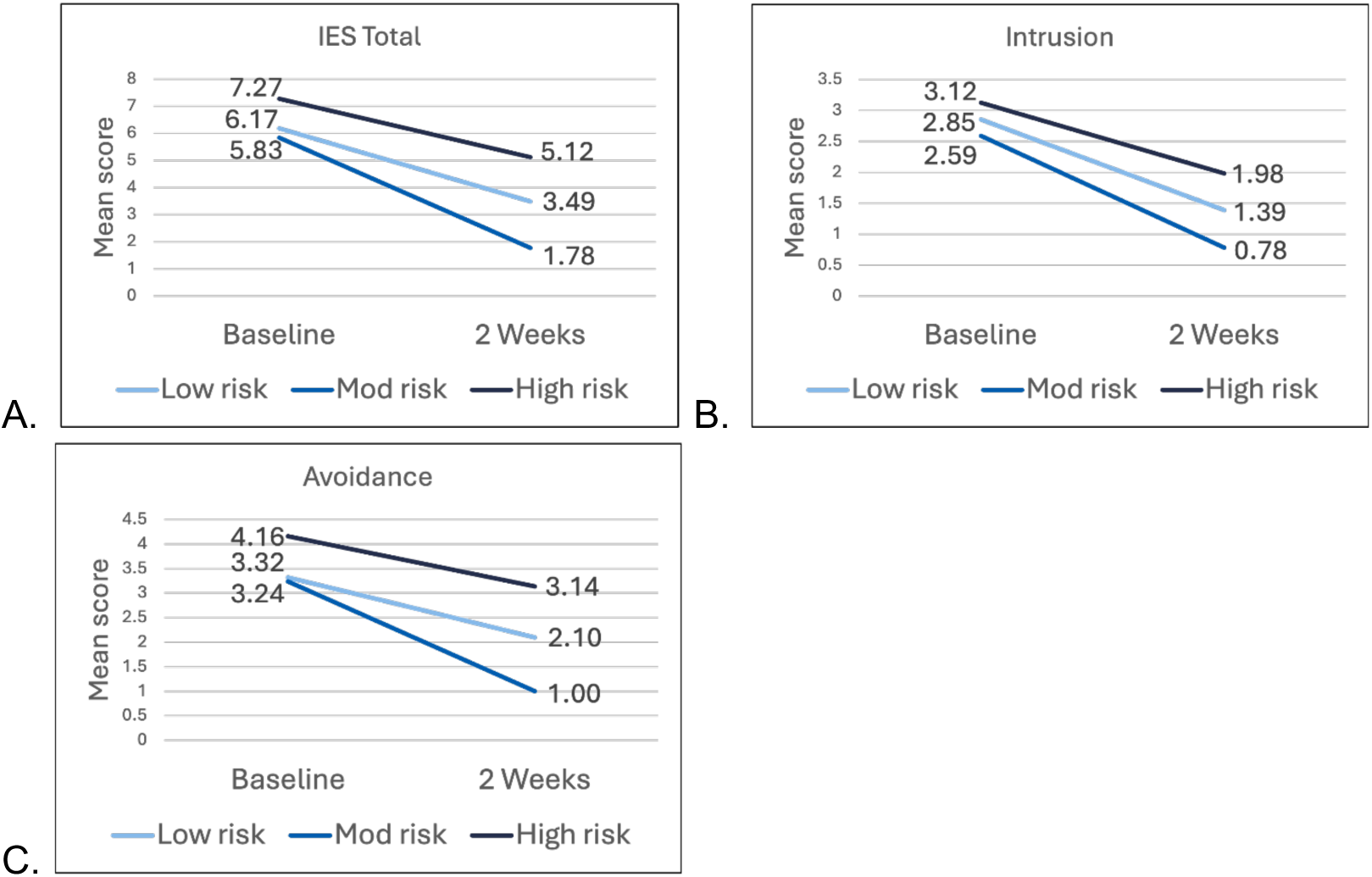
Comparison of mean Impact of Event Scale (IES) scores measuring A) overall glaucoma-related anxiety, B) intrusion and C) avoidance across PRS groups from Baseline to Week 2.

Controlling for baseline IES scores, week 2 total IES scores were associated with PRS (Table 3). This association was stronger for the avoidance subscale (Table 3, Figure 3). Pairwise comparisons showed higher total glaucoma anxiety in the high-risk compared with the moderate-risk group (*t* = –2.36, *p* = .020), while there was not a significant difference between low- and high-risk groups (*t* = –1.79, *p* = .077). The relationship was not significantly changed when controlling for covariates (Supplementary Table 6).

### Test-related distress (FACToR)

Test-related distress was assessed using the FACToR scale which includes four subscales for positive experience, negative emotions, uncertainty, and privacy concerns. Higher scores for each subscale indicate greater psychosocial impact, except for positive experience for which higher scores reflect lower impact. Negative emotions and uncertainty scores were higher, and positive experience scores were lower in the high-risk PRS group compared to the low and moderate PRS groups (Table 4).

**Table 4.** FACToR subscale scores across PRS groups.

| Week 2 scores |  | PRS group effect at Week 2 and pairwise comparisons<sup>1</sup> |  |
| --- | --- | --- | --- |
| Mean (SD) |  | Comparison groups | Test-statistic, p-value) |
| **Negative Emotions (range: 0-12)** |  |  |  |
| All receivers | 0.86 (1.4) | All receivers | $F = 5.70$, .004** |
| PRS (low) | 0.59 (1.1) | High risk v low risk | $t = -2.87$, .005** |
| PRS (moderate) | 0.32 (0.9) | High risk v moderate risk | $t = -4.23$, <.001*** |
| PRS (high) | 1.47 (1.7) | Moderate risk v low risk | $t = 1.29$, .200 |
| **Positive Experience (range: 0-16)** |  |  |  |
| All receivers | 8.17 (3.3) | All receivers | $\chi^2 = 20.80$, <.001*** |
| PRS (low) | 9.15 (3.7) | High risk v low risk | $z = 3.12$, .002** |
| PRS (moderate) | 8.66 (3.2) | High risk v moderate risk | $z = 4.37$, <.001*** |
| PRS (high) | 7.00 (2.8) | Moderate risk v low risk | $z = 1.14$, .254 |

| **Uncertainty (range: 0-12)** |  |  |  |
| --- | --- | --- | --- |
| All receivers | 1.70 (2.2) | All receivers | $X^2 = 11.58, .003^{**}$ |
| PRS (low) | 1.10 (1.5) | High risk v low risk | $z = 2.78, .017^*$ |
| PRS (moderate) | 1.12 (1.5) | High risk v moderate risk | $z = 2.99, .008^{**}$ |
| PRS (high) | 2.62 (2.7) | Moderate risk v low risk | $z = 0.21, 1.00$ |
| **Privacy concerns (range: 0-8)** |  |  |  |
| All receivers | 0.56 (1.1) | All receivers | $X^2 = 4.17, .124$ |
| PRS (low) | 0.29 (0.6) | High risk v low risk | $z = 12.09, .064$ |
| PRS (moderate) | 0.46 (1.0) | High risk v moderate risk | $z = 10.43, .110$ |
| PRS (high) | 0.81 (1.4) | Moderate risk v low risk | $z = 1.66, .809$ |
| **Privacy concerns: health insurance status (range: 0-4)** |  |  |  |
| All receivers | 0.44 (0.85) | All receivers | $X^2 = 4.31, .116$ |
| PRS (low) | 0.24 (0.49) | High risk v low risk | $z = 12.02, .059$ |
| PRS (moderate) | 0.39 (0.86) | High risk v moderate risk | $z = 10.24, .107$ |
| PRS (high) | 0.65 (1.02) | Moderate risk v low risk | $z = 1.78, .790$ |
| **Privacy concerns: employment status (range: 0-4)** |  |  |  |
| All receivers | 0.11 (0.38) | All receivers | $X^2 = 2.56, .278$ |
| PRS (low) | 0.05 (0.22) | High risk v low risk | $z = 6.10, .128$ |
| PRS (moderate) | 0.07 (0.26) | High risk v moderate risk | $z = 4.52, .260$ |
| PRS (high) | 0.20 (0.53) | Moderate risk v low risk | $z = 1.59, .708$ |

<sup>1</sup>Overall group differences were assessed using one-way ANOVA ( $F$ ) or Kruskal–Wallis tests ( $\chi^2$ ), as appropriate.
Asterisks denote statistical significance: \* $p < 0.05$ ; \*\* $p < 0.01$ ; \*\*\* $p < 0.001$

Positive experience was not associated with any of the assessed demographic factors or psychosocial outcomes (Supplementary Table 5), and the relationship between PRS and positive experience remained significant when controlling for covariates (Supplementary Table 6). Sex was associated with negative emotions and uncertainty scores, where women experienced greater levels of negative emotions (mean ranks of women = 71.94 and men = 57.96, *U* = 1596.0, *p* = .020) and uncertainty (mean ranks of women = 72.69 and men = 56.60, *U* = 1532.0, *p* = .015), relative to men (Supplementary Table 5). A positive family history of glaucoma was also associated with greater levels of uncertainty (mean ranks of positive family history = 76.45 and negative or unknown family history = 61.84, *U* = 1577.0, *p* = .027) (Supplementary Table 5). When controlling for these covariates in their respective models, the relationship between PRS and negative emotions, and between PRS and uncertainty, remained significant (Supplementary Table 6).

Younger individuals experienced greater levels of privacy concerns than older individuals (mean ranks: 50-59 years = 71.95, 60-69 years = 72.13, 70-79 years = 67.55, >80 years = 47.00, χ² = 8.65, *p* = .034), while higher educational attainment was associated with lower levels of privacy concerns (mean ranks university = 61.24, vocational/technical = 64.31; high school = 67.94, χ² = 6.05, *p* = .048; Supplementary Table 5). The privacy concerns subscale comprised two items relating to the potential impact of genetic test results on health insurance and employment status. When these items were analysed together, participants receiving high-risk PRS results reported greater concern than those receiving lower-risk results; however, the differences were not statistically significant and overall levels of concern were low. A similar pattern was observed for both items individually, although concerns regarding potential implications for health insurance were greater than concerns relating to employment.

## Discussion

As glaucoma PRS testing becomes increasingly available in clinical practice, understanding its psychosocial impact is critical (17). Our previous work indicated a strong hypothetical level of interest in PRS testing for glaucoma amongst unaffected individuals (71.3%) and individuals with glaucoma (69.4%) (19,29). A recent study by our team evaluated enrolment in the GRADE study among individuals who completed a survey on interest in glaucoma polygenic testing.

Expressed interest was not associated with enrolment, whereas a positive family history and higher education were (29,30). While prior research has examined psychosocial outcomes following PRS testing in conditions including inherited cancers and cardiovascular disease (31–36), this study is the first to explore these outcomes in the context of glaucoma.

### Psychosocial findings

Overall, levels of generalised depression, anxiety, stress, and glaucoma-specific anxiety were low at baseline, and remained low following results disclosure, suggesting that PRS results elicit minimal psychological distress. Across the cohort, small reductions in glaucoma-specific anxiety, general anxiety, stress, and depression were observed after disclosure, consistent with prior studies (32,37). This pattern may reflect a reduction in anticipatory anxiety once results are received (38).

General measures of depression, anxiety, and stress were not moderated by PRS group or demographic variables, including sex, education, family history, and result expectations. However, despite low absolute scores, participants in the high-risk PRS group reported higher glaucoma anxiety compared to those in the moderate-risk group. Similarly, test-related distress showed some impact following disclosure, with high-risk participants reporting greater negative emotions and fewer positive experiences than moderate- and low-risk participants, although scores remained within normal-to-mild ranges.

These findings are consistent with the limited literature examining psychosocial outcomes following PRS disclosure for other common conditions. Studies in breast cancer and melanoma reported low overall psychological distress, with modestly higher distress or worry among individuals receiving higher-risk results (31,32,37). Importantly, these differences have not been associated with clinically significant distress, reinforcing the general psychological safety of PRS testing when delivered with appropriate support.

### Recall, comprehension of results, and knowledge about PRS results

Two weeks after receiving results, recall was high, with 78% of participants accurately recalling their personal risk relative to the population. Participants were not provided with a numerical PRS, but instead received a categorical result (high, average, or low risk relative to the population). This approach was informed by our previous work, which found that even highly educated participants found PRS percentiles confusing and sometimes misinterpreted them as absolute risk, leading to unnecessary worry (23). Other studies similarly emphasise the importance of contextualising genetic risk relative to population norms (39,40).

Almost one in eight participants reported a mismatch between their reported family history and PRS result. Family history and PRS are complementary but independent measures across a range of diseases including glaucoma (41,42). Subjective perceptions of risk based on experiential knowledge such as family history however, can be resistant to change, with persistence of inaccurate or exaggerated perceptions observed even after genetic counselling and testing (43–45). While breast cancer PRS studies have reported closer alignment between self-perceived and clinically assessed risk, both before and after testing (46,47) family history of glaucoma is frequently misunderstood or misreported, which could account for our different results (48–50).

Knowledge of glaucoma and PRS was generally poor, although knowledge of glaucoma was higher than genetic concepts. Knowledge improved following receipt of results, but only modestly. This highlights a need for improved pre- and post-test education and counselling to help participants understand and contextualize their results, which may present challenges for broader implementation. Future research should assess whether recall and comprehension are sustained over longer follow-up and whether this translates into changes in screening behaviour, as well as identify scalable strategies to improve education and understanding. This challenge is not unique to ophthalmology and is observed across multiple disciplines. Current genetic counselling capacity is insufficient to meet growing demand. To improve efficiency and reduce pressure on specialist genetics services, expanding access to genomic testing will require mainstreaming, whereby aspects of testing would be undertaken by clinicians without primary training in genetics (51–53).

### Privacy concerns

Participants reported low levels of privacy concerns. Participants indicated slightly higher concern about their results impacting their health insurance status than their employment status. Although fear of employment and insurance discrimination is a well-described barrier to genetic testing (54–56), most participants in this study were aged over 50 and identified as retired, making concerns about workplace discrimination less personally relevant in this cohort, as reflected in the study findings.

While not statistically significant, individuals with a high-risk PRS were more concerned about potential implications for their health insurance compared with those in the low-risk group. In Australia, private health insurance is not based on a risk assessment of health and, as such, insurers cannot use genetic test results to deny cover or increase premiums (57). However, participants may be conflating potential implications for health insurance with those for life insurance, which operates differently. A current industry-led moratorium restricts the use of genetic information by life insurers; however, there is limited guidance regarding the specific implications for PRS. The insurance-related concerns reported by participants may therefore reflect perceived implications for future life insurance applications. This finding suggests that clear counselling regarding data protection and the insurance implications of genetic testing is important. On April 1 2026, *The Genetic Testing Protections in Life Insurance and Other Measures Bill* (58) was passed by both houses of the Australian Federal Parliament (59) and will come into effect in October 2026 (60), which may help reduce misunderstanding and concern for future testing.

### Decisional regret and non-participation

Decisional regret was low across all PRS groups, suggesting participants were comfortable with their decision to receive results. However, only 46.5% of invited participants from the GRADE study consented to result disclosure, indicating a potential self-selection bias. Compared with non-participants (including those who actively declined or could not be contacted), those who enrolled were younger and more likely to be female and to have glaucoma or a family history of the condition, suggesting that individuals with these characteristics may be more motivated to receive genetic risk information. This may explain the higher percentage of women in the high-risk PRS group in this study compared to the moderate and low risk PRS groups despite higher rates of primary open-angle glaucoma being described in men (61,62).

Reasons for non-participation were not systematically collected. However, limited time availability, as well as medical factors (including personal or caregiving responsibilities), and reduced cognitive capacity to complete surveys were self-reported; factors that may be more common with advanced age. Whilst age and life experience have been proposed as a source of resilience when receiving genetic results (63), age has been associated with lower uptake of genetic testing (64). Whilst there is limited research regarding the reasons older adults engage at a lower rate with genetic testing, previously cited reasons include concern for negative psychological outcomes, lack of perceived utility, scepticism about genetic testing, concern regarding privacy and confidentiality, concerns regarding validity, and time limitations. (65,66)

### Limitations

The cohort was primarily European and English-speaking and so findings may not be generalised across populations. It is notable that PRS performance is lower in non-European populations, and these groups may experience greater marginalization in healthcare (67). With the Australian population representing diverse cultural backgrounds, psychosocial outcomes will need to be assessed in other populations for generalisability. However, PRS that perform equally across ancestral groups are also needed to accurately assess this. Participants were self-selected and highly educated, and under half of those invited consented to receive their results. The enrolment may have biased the cohort toward individuals with lower psychological sensitivity to receiving genetic risk information. There is a need for further research into the motivations of individuals who declined participation to assess whether they may encounter high adverse outcomes and require further support.

## Conclusions

This study supports the feasibility of glaucoma polygenic testing with minimal psychological harm. The higher negative emotions and lower positive experiences observed among high-risk participants may reflect the absence of clear clinical guidance for follow-up and management, underscoring the need for evidence-based practice recommendations. Development of clinical guidelines for managing high-risk PRS results should therefore be prioritised. Modest knowledge of glaucoma and genetics emphasises the need for targeted education to improve understanding and contextualisation of PRS results. Future studies are warranted to assess sustained psychological outcomes, recall and comprehension of results, and potential behavioural responses to PRS disclosure.

## Data Availability

All data produced in the present study are available upon reasonable request to the authors.

