## Supplementary Material for "Short-term psychosocial outcomes following disclosure of glaucoma polygenic risk score, INSiGHT Study"

### Supplementary Information

**Supplementary Table 1.** Internal consistency scores across measures

| Psychosocial | Baseline ω | Week 2 ω |
|---|---|---|
| Genetic determinism (removing item 2) | .695 | N/A |
| Decisional regret | N/A | .88 |
| DASS - Depression Subscale | .904 | .87 |
| DASS - Stress Subscale | .856 | .87 |
| DASS - Anxiety Subscale | .776 | .56 |
| Overall IES scores (glaucoma anxiety) | .951 | .93 |
| IES Intrusion subscale | .912 | .92 |
| IES Avoidance subscale | .908 | .82 |
| FACToR - Negative Emotions Subscale | N/A | .67 |
| FACToR - Positive Experience Subscale | N/A | .68 |
| FACToR - Uncertainty Subscale | N/A | .86 |
| FACToR - Privacy concerns | N/A | .51 |

DASS = Depression, Anxiety and Stress scale, IES = Impact of Events scale as a measure of glaucoma-specific anxiety, FACToR = Feelings About genomiC Testing REsults scale as a measure of test-related distress. Internal consistency is reported as McDonald's Omega where possible and Cronbach's alpha for Privacy Concerns FACToR subscale where use of McDonald's Omega was precluded due to there being fewer than items. Decisional regret and the FACToR scale were not measured at Baseline.

**Supplementary Table 2.** Result expectation and recall

| Participant response | Low risk (n=41) | Moderate risk (n=41) | High risk (n=51) |
|---|---|---|---|
| **Perceived concordance between PRS result and family history** | | | |
| Yes, my result fit with my family history | 21 (51.2%) | 20 (48.8%) | 24 (47.1%) |
| No, my result didn't fit with my family history | 7 (17.1%) | 2 (4.9%) | 9 (17.6%) |
| I was unsure what result to expect based on my family history | 13 (31.7%) | 19 (46.3%) | 18 (35.3%) |
| **Accuracy of PRS result recall** | | | |
| Correctly recalled their results | 31 (75.6%) | 28 (68.3%) | 45 (88.2%) |
| Recalled their results as higher than actual | 6 (14.6%) | 1 (2.4%) | N/A |
| Recalled their results as lower than actual | N/A | 8 (19.5%) | 4 (7.8%) |
| Did not recall their results | 4 (9.8%) | 4 (9.8%) | 2 (4.9%) |

**Supplementary Table 3.** Result expectation between PRS groups alignment with family history

| Outcome (vs. "Unsure") | Predictor Comparison | B | p-value |
|---|---|---|---|
| Expectation aligning with FH | Low vs. High PRS | 0.22 | 0.648 |
| | Moderate vs. High PRS | -0.21 | 0.645 |
| | Family history | -0.15 | 0.721 |
| Expectation not aligning with FH | Low vs. High PRS | 0.15 | 0.812 |
| | Moderate vs. High PRS | -1.48 | 0.085 |
| | Family history | -0.42 | 0.468 |

**Supplementary Table 4.** Questions for the knowledge measure including comparison of proportion correctly answered at baseline compared to week 2

| Question | Correct baseline response (n, %) | Correct week 2 response (n, %) |
|---|---|---|
| All individuals at high risk for glaucoma will develop glaucoma (F) | 44 (33.1) | 60 (45.1) |
| The interpretation of a high or low risk is the same for everyone (F) | 52 (39.1) | 63 (47.4) |
| Common risk variants associated with glaucoma risk also increases a person's risk for other eye diseases (F) | 12 (9.0) | 16 (12.0) |
| There is more than one DNA change that can increase a person's risk for glaucoma (T) | 26 (19.5) | 41 (30.8) |
| It is possible to be diagnosed with glaucoma solely due to chance (T) | 70 (52.6) | 83 (62.5) |
| Most individuals who develop glaucoma do not have family history of the disease (T) | 7 (5.3) | 16 (12.0) |
| A person inherits DNA changes associated with glaucoma risk from both parents (T) | 24 (18.0) | 34 (25.6) |
| A person may be at increased risk for glaucoma if they have several close relatives with glaucoma (T) | 109 (82.0) | 111 (83.5) |
| If a person has a high risk for glaucoma there are screening and preventative options available (T) | 108 (81.2) | 124 (93.2) |
| An eye test can only detect glaucoma when it is more advanced (F) | 85 (63.9) | 93 (69.9) |
| Early detection of glaucoma means a | 121 (91.0) | 122 (91.7) |

| | | |
|---|---|---|
| greater chance of not losing vision (T) | | |
| A person with early glaucoma may not be aware that they have the disease (T) | 126 (94.7) | 128 (96.2) |

n = 133 participants who completed Baseline and Week 2 surveys. True (T) and False (F) refer to the correct answer for each question.

**Supplementary Table 5.** Relationship between demographic factors and psychosocial outcomes

| Measure | Demographic factor | Test statistic | P value |
|---|---|---|---|
| **Decisional regret** | Sex | *U* = 2124.50 | .609 |
| | Education | *H* = 2.02 | .365 |
| | Family History | *U* = 2182.50 | .425 |
| | Age | *H* = 4.42 | .219 |
| **Week 2 Depression^** | Sex | *F* = 0.32 | .574 |
| | Education | *F* = 0.57 | .567 |
| | Family History | *F* = 2.60 | .109 |
| | Age | *F* = 0.06 | .980 |
| **Week 2 Anxiety^** | Sex | *F* = 0.42 | .517 |
| | Education | *F* = 0.77 | .470 |
| | Family History | *F* < 0.01 | .949 |
| | Age | *F* = 0.33 | .808 |
| **Week 2 Stress^** | Sex | *F* < .001 | .988 |
| | Education | *F* = 1.81 | .168 |
| | Family History | *F* = 1.12 | .291 |
| | Age | *F* = 0.51 | .675 |
| **Week 2 IES Total^** | Sex | *F* = 0.21 | .648 |
| | Education | *F* = 0.66 | .517 |
| | Family history | *F* = 1.11 | .295 |
| | Age | *F* = 0.50 | .686 |
| **Week 2 IES Intrusion^** | Sex | *F* < .01 | .941 |
| | Education | *F* = 0.92 | .400 |
| | Family history | *F* = 0.10 | .758 |
| | Age | *F* = 0.74 | .528 |
| **Week 2 IES Avoidance^** | Sex | *F* = 0.04 | .843 |
| | Education | *F* = 0.55 | .576 |
| | Family history | *F* = 1.89 | .171 |
| | Age | *F* = 0.26 | .856 |
| **Week 2 FACToR positive experience** | Sex | *U* = 1872.50 | .482 |
| | Education | H = 1.79 | .410 |
| | Family history | U = 1971.50 | .815 |
| | Age | H = 0.77 | .856 |
| **Week 2 FACToR negative emotions** | Sex | *U* = 1596.00 | .020* |
| | Education | *H* = 1.26 | .532 |

| | | | |
|---|---|---|---|
| | Family history | *U* = 2029.50 | .963 |
| | Age | *H* = 3.67 | .299 |
| **Week 2 FACToR uncertainty** | Sex | *U* = 1532.00 | .015* |
| | Education | *H* = 1.86 | .395 |
| | Family history | *U* = 1577.00 | .027* |
| | Age | *H* = 3.93 | .270 |
| **Week 2 FACToR privacy concerns** | Sex | *U* = 1721.00 | .080 |
| | Education | *H* = 6.05 | .048* |
| | Family history | *U* = 1942.00 | .645 |
| | Age | *H* = 8.65 | .034* |

^ = controlling for baseline scores

Asterisks denote statistical significance: * p<0.05; ** p < 0.01; *** p<0.001

**Supplementary Table 6.** Effect PRS on psychosocial outcomes controlling for demographic or psychosocial factors.

| Scale scores | Demographic/psychosocial factor | F-statistic | P value |
|---|---|---|---|
| **Decisional regret** | Sex | 0.11 | .894 |
| | Education | 0.10 | .910 |
| | Family History | 0.10 | .907 |
| | Age | 0.16 | .855 |
| | Knowledge | 0.18 | .836 |
| | Genetic Determinism | 0.20 | .819 |
| | Result Expectation | 0.15 | .864 |
| **Week 2 Depression^** | Sex | 1.66 | .194 |
| | Education | 1.69 | .189 |
| | Family History | 2.01 | .138 |
| | Age | 1.76 | .176 |
| | Knowledge | 1.77 | .174 |
| | Genetic Determinism | 1.88 | .157 |
| | Result Expectation | 1.77 | .175 |
| **Week 2 Anxiety^** | Sex | 0.02 | .986 |
| | Education | 0.03 | .969 |
| | Family History | 0.02 | .981 |
| | Age | 0.03 | .970 |
| | Knowledge | 0.02 | .980 |
| | Genetic Determinism | 0.05 | .948 |
| | Result Expectation | 0.03 | .971 |
| **Week 2 Stress^** | Sex | 0.85 | .430 |
| | Education | 0.72 | .490 |
| | Family History | 0.83 | .441 |
| | Age | 0.84 | .436 |
| | Knowledge | 0.79 | .454 |
| | Genetic Determinism | 0.91 | .407 |
| | Result Expectation | 0.88 | .419 |
| **Week 2 IES Total^** | Sex | 2.82 | .063 |

| | | | |
|---|---|---|---|
| | Education | 3.27 | .041* |
| | Family history | 2.62 | .077 |
| | Age | 3.17 | .045* |
| | Knowledge | 3.08 | .050 |
| | Genetic Determinism | 3.07 | .050 |
| | Result Expectation | 3.18 | .045* |
| **Week 2 IES Intrusion^** | Sex | 0.56 | .572 |
| | Education | 0.61 | .545 |
| | Family history | 0.60 | .551 |
| | Age | 0.57 | .566 |
| | Knowledge | 0.33 | .721 |
| | Genetic Determinism | 0.55 | .577 |
| | Result Expectation | 0.65 | .522 |
| **Week 2 IES Avoidance^** | Sex | 2.51 | .085 |
| | Education | 2.76 | .067 |
| | Family history | 2.06 | .131 |
| | Age | 2.67 | .073 |
| | Knowledge | 1.80 | .170 |
| | Genetic Determinism | 2.59 | .079 |
| | Result Expectation | 2.66 | .074 |
| **Week 2 FACToR positive experience** | Sex | 4.20 | .017* |
| | Education | 5.04 | .008** |
| | Family history | 5.21 | .007** |
| | Age | 6.92 | .001** |
| | Knowledge | 1.43 | .243 |
| | Genetic Determinism | 0.37 | .693 |
| | Result Expectation | 5.24 | .007** |
| **Week 2 FACToR negative emotions** | Sex | 9.23 | <.001*** |
| | Education | 12.31 | <.001*** |
| | Family history | 12.22 | <.001*** |
| | Age | 11.73 | <.001*** |
| | Knowledge | 11.82 | <.001*** |
| | Genetic Determinism | 12.11 | <.001*** |
| | Result Expectation | 12.50 | <.001*** |
| **Week 2 FACToR uncertainty** | Sex | 4.21 | 0.17 |
| | Education | 6.25 | .003** |
| | Family history | 4.86 | .009** |
| | Age | 6.11 | .003** |
| | Knowledge | 6.03 | .003** |
| | Genetic Determinism | 6.25 | .003** |
| | Result Expectation | 6.43 | .002** |
| **Week 2 FACToR privacy concerns** | Sex | 1.31 | .272 |
| | Education | 2.19 | .116 |
| | Family history | 1.94 | .148 |
| | Age | 2.57 | .080 |
| | Knowledge | 1.84 | .163 |
| | Genetic Determinism | 2.14 | .122 |

| | | | |
|---|---|---|---|
| | Result Expectation | 2.12 | .125 |

^When controlling for baseline scores

Asterisks denote statistical significance: * $p<0.05$; ** $p < 0.01$; *** $p<0.001$
